# Treatment Outcome of Adults Receiving Virtual Cognitive Behavior Therapy for an Eating Disorder

**DOI:** 10.1101/2025.04.07.25325397

**Authors:** Jessica H. Baker, Nickolas M. Jones, David Freestone, Lara Effland, Cara Bohon

## Abstract

**Objective:** The aim of this paper is to evaluate the effectiveness of enhanced cognitive behavioral therapy (CBT-E) adapted to be delivered via telehealth in a real-world, clinic treatment setting by a multi-disciplinary team for adults with an eating disorder.

**Method:** A retrospective analysis of treatment outcomes was conducted on adult patients (18+) who received adapted CBT-E, a transdiagnostic treatment approach specifically modified for the treatment of eating disorders. Outcome included weight and eating disorder, depression, and anxiety symptoms. Survival analyses were used to assess length of stay, weight restoration and alleviation of eating disorder, depression, and anxiety symptoms; multilevel models assessed outcome trajectories over treatment time.

**Results:** The patient sample (n = 1,718) was predominantly white (73%), cisgender women (86%), with a mean age of 30 (SD = 12). Diagnoses included anorexia nervosa (AN, 56%), binge-eating disorder (BED, 24%), bulimia nervosa (BN, 7%), and other specified feeding and eating disorder (OSFED, 11%). Approximately 51% of patients with weight restoration targets achieved weight restoration (95% of their target weight) by week 40 of treatment. By week 40 in treatment, 49% of patients reached subclinical levels on the EDE-Q, 58% on the PHQ-8, and 56% on the GAD-7.

**Discussion:** Adapted CBT-E delivered via telehealth by a multi-disciplinary team is effective in improving transdiagnostic eating disorder symptoms, depression, and anxiety in an outpatient setting. Outcomes were consistent across diagnoses, demonstrating the feasibility and effectiveness of virtual CBT-E. However, variability in treatment length makes direct comparison with clinical trial end-of-treatment outcomes challenging.

**Public Health Significance:** This study shows that virtually-delivered CBT-E effectively treats eating disorders by improving symptoms and accessibility to treatment. Expanding virtual treatment can significantly reduce barriers to care, reaching more individuals and lessening the public health impact of these disorders.

## Introduction

Prevailing evidence suggests that cognitive behavioral therapy (CBT) is an effective treatment for eating disorders (de Jong, Schoorl, & Hoek 2018; Linardon, Wade, de la Piedad Garcia, & Brennan, 2017; Dahlenburg, Gleaves, & Hutchinson, 2019). Enhanced CBT (CBT-E) is a variant of CBT modified specifically for the treatment of eating disorders (Fairburn 2008). CBT-E was developed to be transdiagnostic such that it involves targeting the maintaining mechanism(s) of the eating disorder rather than a specific diagnostic category (Fairburn, 2008). Although clinical trials have shown CBT-E to be generally effective (Atwood & Friedman, 2020), less is known about the effectiveness in real world treatment settings (Ebrahimi et al., 2024) or about delivery by multidisciplinary teams, despite some work in collaborative and integrated settings (Bray et al., 2023). Furthermore, with the rise of telehealth there is a need to evaluate the effectiveness of CBT-E via telemedicine.

Systematic reviews and meta-analyses indicate that CBT-E is generally considered the first line outpatient treatment for eating disorders (de Jong, Schoorl, & Hoek, 2018; Duggan, Hardy, & Waller, 2025; Linardon, Wade, de la Piedad Garcia, & Brennan, 2017; Dahlenburg, Gleaves, & Hutchinson, 2019; Öst et al., 2023). Although clinical trials have been useful in establishing the effectiveness of CBT-E, they often have limited external validity. The treatment plan, patient sample, and criteria for success in clinical trials have been stringently specified *a priori*. Indeed, clinical trials have strict inclusion and exclusion criteria often excluding certain comorbidities (e.g., suicidality, substance use disorder) and psychosocial circumstances (e.g., unhoused, inability to commit to trial duration), use of certain psychiatric medications, medical instability, and previous treatment (de Jong et al., 2016). This results in a patient population that is not representative of the typical patient seeking care for an eating disorder. To bridge this gap, real world data is needed to understand how eating disorder treatment delivered via CBT-E performs in the broader patient population.

While the one-year prevalence of eating disorders (Galmiche et al., 2019) are comparable to other common mental health illnesses (National Mental Health Alliance, 2021), access to treatment is worse (Reinert et al., 2021). Approximately 80% of people with an eating disorder never receive care (Hart et al., 2011). There are myriad reasons for the low rates of access including: geographic barriers, insurance coverage, high out-of-pocket costs, training required for CBT-E, and long wait lists to receive evidence-based care (Ali et al., 2017; Attia et al., 2016; Cachelin et al., 2001; Duffy et al., 2016; Innes et al., 2017; Veillette et al., 2018; Murray & Le Grange, 2014). The desire for telemedicine has risen significantly in recent years (Barnett et al., 2018), and its availability increases access to evidence-based care that may not be available locally to people who need it. To date, research has shown that treatments provided virtually are similarly effective to in-person care (Levinson, Spoor, Keshishian, & Pruitt, 2021; Anderson, Byrne, Crosby, & Le Grange, 2017). However, given the challenges that may exist with telehealth (e.g., inability to view body language, in-home distractions, technical difficulties), it is essential to evaluate the effectiveness of evidence-based in-person care when delivered in a new manner.

Taken together, the aim of this paper is to evaluate the effectiveness of CBT-E delivered via telehealth by a multidisciplinary team in a clinical treatment setting. Thus, we describe the CBT-E treatment approach, report treatment outcomes, and discuss challenges translating clinical trials to real-world settings.

## Method

### Patients

Patients were enrolled in a virtual eating disorder treatment program that treats patients aged 6+. The current study includes adult patients (18+) enrolled in the program from August 2021 to December of 2024. Patients could be discharged or currently receiving care. Sessions were conducted via a HIPAA compliant telehealth platform. The evaluation of our patient treatment outcomes was reviewed by the Western Institutational Review Board (WIRB). It was determined evaluation of patient treatment outcomes do not meet the definition of human subjects research and are therefore considered exempt from IRB oversight.

### Treatment Overview

Adult patients diagnosed with anorexia nervosa (AN), bulimia nervosa (BN), binge-eating disorder (BED), and other specified feeding and eating disorder (OSFED) received CBT-E as first-line treatment. The structure of the treatment was delineated into four stages: 1) engaging the patient in the treatment and building motivation for change through rapport and trust with the team and the treatment; 2) a bridge stage for the treatment team to review the patient’s progress in stage one; 3) the third stage was the core of CBT-E aiming to target maintaining factors of the patient’s eating disorder (Fairburn 2008). Each patient worked through one of five modules that pertain to their specific eating disorder symptoms; 4) ending treatment and relapse prevention (Fairburn 2008). A step-down team was identified and scheduled as needed for the patient to continue longer-term treatment support to sustain recovery.

The multidisciplinary patient care team included a therapist, dietitian, peer mentor, and medical provider as needed. The peer mentor has recovered from an eating disorder and provided a source of hope and motivation for the patient to see that recovery is possible. The treatment was conducted via weekly individual sessions with dietitian and/or therapist. Licensed providers received approximately10 hours of training and 12 hours of consultation post-training. Therapists continued with clinical supervision and consultation via weekly individual and group supervision. The CBT-E program was modified slightly for virtual care as well as for inclusivity. Specifically, all language was modified for weight inclusivity such that weight was masked unless deemed clinically appropriate, stage 1 included a support person if the patient had a support available, and stage 3 treatment was divided between the therapist and dietitian. The division between treatment team members focused sessions on the provider’s scope and expertise. Broad modules were offered via groups to ease the training burden for team members and aid in implementation of evidence-based treatment strategies (e.g., perfectionism, core low self-esteem, emotion regulation skills).

### Study Design and Overview

To evaluate treatment effectiveness, we collected various outcomes throughout the course of treatment. Measures are completed as part of standard care.

### Measures

#### Weight

Weight was collected using BodyTrace Scale, a connected device that automatically sends weight data electronically to the electronic medical record, starting in March 2024. Prior to this, patients completed weight collection via in-person appointments at primary care or student health centers or self-report weight from a scale at home. Patients were instructed to check weight twice weekly with minimal clothing, after voiding, and prior to food or beverage consumption. For patients needing weight restoration, target weight was determined by the registered dietitian using Centers of Disease Control age-adjusted body mass index growth charts and the patient’s individual growth trajectory from historical medical records (Kuczmarski et al., 2002). Weight restoration here was defined as achieving 95% of target weight set by a practitioner at the onset of treatment. Analyses evaluating weight change only included patients with a weight restoration treatment goal.

#### Eating disorder symptoms

To assess eating disorder symptoms, patients completed the Eating Disorder Examination-Questionnaire (EDE-Q) monthly for the first 4 months of treatment and quarterly thereafter. The EDE-Q has 28 items and is made up of 4 subscales (restraint, eating concerns, weight concerns, and shape concerns) and a global score. A global score greater than 2.8 indicates clinically significant eating disorder symptoms (Mond et al., 2008). Internal consistency was acceptable (Cronbach’s α = .94).

#### Depression

Patients completed the eight-item Patient Health Questionnaire (PHQ-8) (Kroenke et al., 2009; Razykov, Ziegelstein, Whooley, & Thombs 2012) monthly for the first 4 months of treatment and quarterly thereafter. The PHQ-8 reflects the Diagnostic and Statistical Manual of Mental Disorders diagnostic criteria for depression and is a valid measure for depression across diverse populations (Huang, Chung, Kroenke, Delucchi, & Spitzer, 2006). The PHQ-8 asks the frequency of depressive symptoms within the past two weeks. Response options range from “not at all” to “nearly every day.” Total scores range from zero to 24. Scores of 10 and above reflect moderate to severe depression. Internal consistency was acceptable (Cronbach’s α = .86).

#### Anxiety

Patients completed the seven-item Generalized Anxiety Disorder Questionnaire (GAD-7; Spitzer, Kroenke, Williams, & Lowe, 2006) monthly for the first 4 months of treatment and quarterly thereafter. The GAD-7 reflects diagnostic criteria for generalized anxiety disorder. The GAD-7 asks the frequency of anxiety symptoms within the past two weeks. Response options range from “not at all” to “nearly every day.” Total scores range from zero to 21. Scores of 10 and above reflect moderate anxiety (Cronbach’s α = .89).

### Analytic Strategy

Descriptive analyses were conducted to characterize the patient sample. Survival analyses were used to determine: 1) the median length of stay in treatment, 2) the percentage of patients meeting weight restoration targets, and 3) the percentage of patients who achieved sub-clinical status on eating disorder, anxiety, and depression symptom scales over treatment time.

Outcome measurements over time were nested within patients. Unlike clinical trials which typically have a specified treatment end point, treatment length among patients in this real-world sample was variable. Thus, we modeled outcomes through one year of treatment. For comparison, we also modeled treatment outcomes at weeks 20 and 40 as these timepoints generally align with CBT-E clinical trial endpoints (Fairburn et al., 2009; Fairburn et al., 2013). In order to provide accurate estimates of these outcomes at specific weeks of treatment, we fit outcome trajectories using a series of multilevel models. Each model took the following form: outcome_w,j_ ∼ β_0_+ β_0j_ + β_1_log(*w*) + β_1j_log(*w*) + β_2_age + β_3_[age × log(*w*)] + β_4_gender + β_5_[gender × log(*w*)] + β_6_diagnosis + β_7_[diagnosis × log(*w*)]; where *w* represents the treatment week (we took the log because treatment progresses logarithmically over time; Steinberg et al, 2022; Howard et al., 1986) and j indexes the patient (so that β_0j_ and β_1j_ represent random intercepts and slopes).

The random effects account for different patient-level starting points and outcome trajectories over time.

We report unstandardized results using all available data for the full sample and by patient diagnosis. However, the aim of this paper is not to compare the effectiveness of CBT-E across eating disorder diagnoses so results are shown by diagnosis for descriptive purposes only.

All analyses were performed in R (R Core Team, 2024; version 4.4.2) using the tidyverse (Wickham et al., 2019; version 2.0.0). Survival analyses were performed using the survival (Therneau, 2024; version 3.8-3) and survminer (Kassambara et al., 2024; version 0.5.0) packages; multi-level modeling was performed using lme4 (Bates et al., 2015; version 1.1-35.5), ggeffects (Lüdecke, 2018; version 2.0.0), and emmeans (Lenth, 2024; version 1.10.6). Project data management and workflow was managed using duckdb (Mühleisen & Raasveldt, 2024; version 1.1.3-1) and targets (Landau, 2021; version 1.9.1).

#### Missingness

Different analyses presented below may have slightly different analytical sample sizes because some patients completed more survey responses than others, and because some patients were missing information necessary for one analysis but not another. Patients were more likely to complete outcome surveys (i.e., EDE-Q, GAD-7, PHQ-8) early in treatment compared to later (all ps < .001). Older adult patients were slightly more likely to fill out the surveys compared with younger adults (all ps < .001). Survey completion for these outcomes was not related to any other variable used here, including gender, diagnosis, and the severity of scores over treatment time. Older adults were more likely to provide weight measurements (*p* < .001); however, missingness on weight was not related to a patient’s overall weight during treatment.

We report complete-case analysis here, but using imputation methods (via the mice package in R; van Buuren & Groothius-Oudshoorn, 2011) gave the same pattern of results, and nearly identical estimated coefficients.

## Results

### Patient characteristics

Patient demographics are shown in Table 1. The patient sample was predominantly cisgender women (n = 1,483; 86%) and white (n = 1,242; 73%). The average patient age was 30 years (*SD* = 12). Approximately 56% of patients were diagnosed with AN, 23% with BED, 7% with BN, and 11% with OSFED. A little over one-third of patients (35%) self-reported receiving treatment at a higher level of care facility (e.g., residential) at some point before the current treatment. Patient characteristics by diagnosis are presented in Supplement Table S1.

**Table 1.**
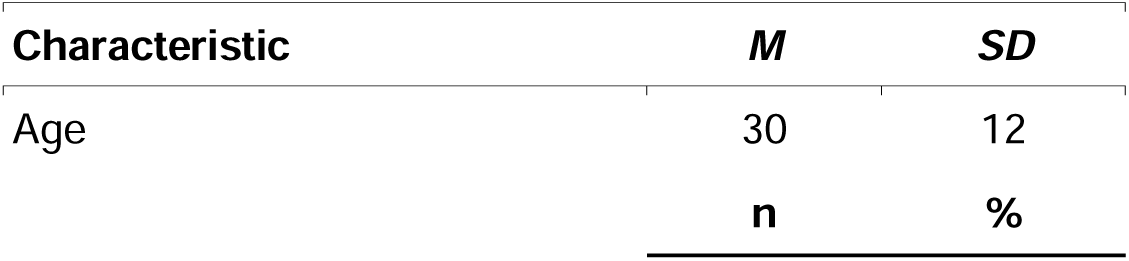
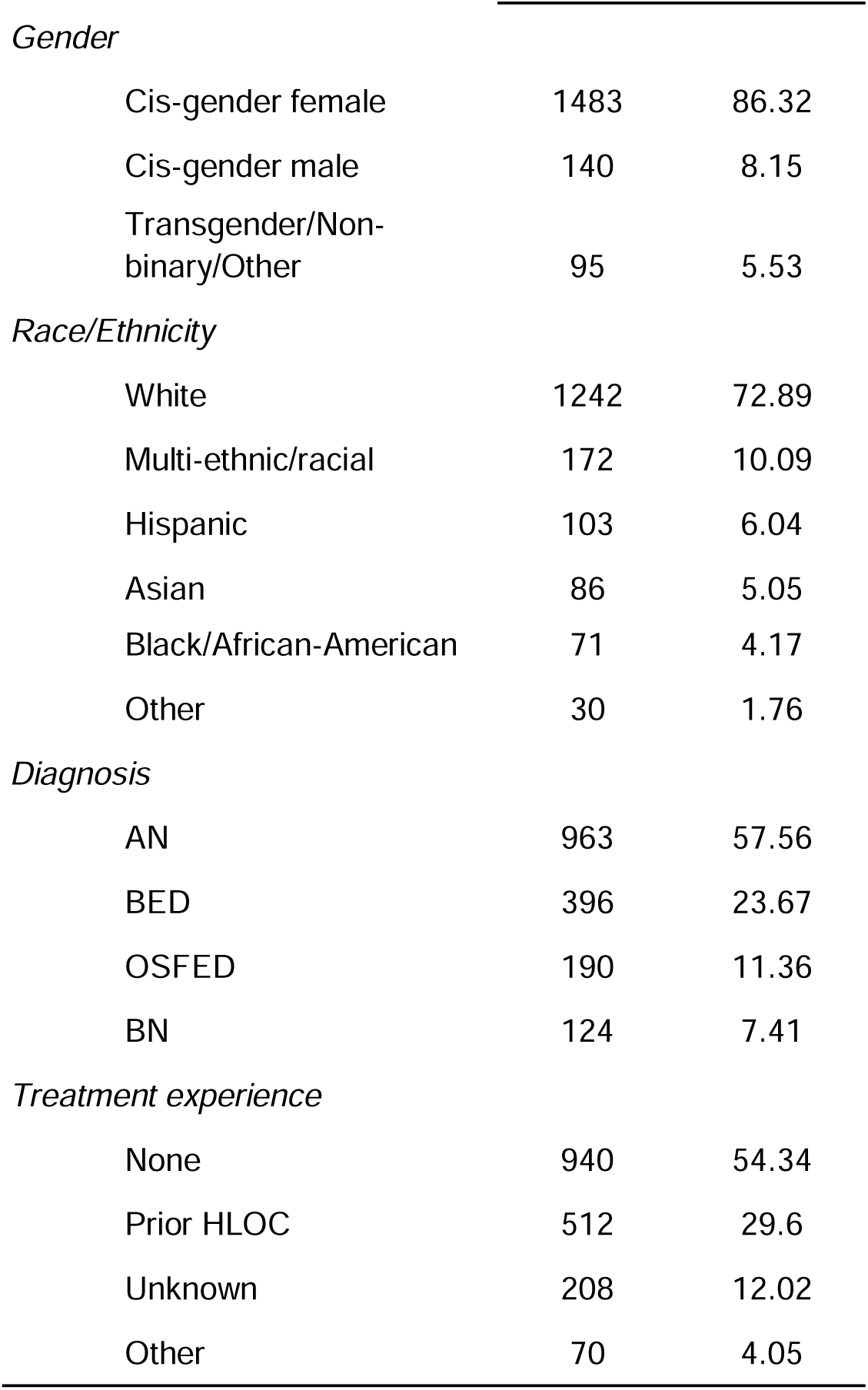
Patient sample characteristics.

| Characteristic | *M* | *SD* |
| --- | --- | --- |
| Age | 30 | 12 |
|  | **n** | **%** |
| *Gender* |  |  |
| Cis-gender female | 1483 | 86.32 |
| Cis-gender male | 140 | 8.15 |
| Transgender/Non-binary/Other | 95 | 5.53 |
| *Race/Ethnicity* |  |  |
| White | 1242 | 72.89 |
| Multi-ethnic/racial | 172 | 10.09 |
| Hispanic | 103 | 6.04 |
| Asian | 86 | 5.05 |
| Black/African-American | 71 | 4.17 |
| Other | 30 | 1.76 |
| *Diagnosis* |  |  |
| AN | 963 | 57.56 |
| BED | 396 | 23.67 |
| OSFED | 190 | 11.36 |
| BN | 124 | 7.41 |
| *Treatment experience* |  |  |
| None | 940 | 54.34 |
| Prior HLOC | 512 | 29.6 |
| Unknown | 208 | 12.02 |
| Other | 70 | 4.05 |

The overall median length of stay was 25 weeks (CI: [22, 29]). Length of stay was significantly longer for patients with OSFED (44 weeks, CI: [30, 52]) relative to all other diagnoses. In treatment, patients were primarily engaged with therapists (41% of sessions) and dietitians (31% of sessions); other less frequent sessions included meeting with a physician, a psychiatrist (if needed), and peer and family mentors. At the onset of treatment, the median number of weekly sessions was approximately three (range: 1 to 7; usually a therapist appointment, dietitian appointment, and one other type of appointment); the median weekly sessions tapered over treatment, down to roughly 2 sessions per week (range: 1 to 6) by week 40.

Approximately 18% (n = 327) of patients required weight restoration and had a target weight set by the dietitian. The median weight gain needed in order to reach target weight was 16lbs. On average, just over half of patients (51%; CI: [42.63, 58.58]) reached weight restoration (95% of target weight) before week 40 of treatment.

Unstandardized multilevel model results for each outcome are presented in Table 2. There was a significant negative relationship between age and anxiety symptoms, suggesting that older patients reported less anxiety than their younger counterparts. Relative to women, men reported significantly less anxiety at the onset of treatment. Moreover, patients identifying as transgender, non-binary, or other reported significantly higher eating disorder, anxiety, and depression symptoms at the onset of treatment relative to women. Patients with BED reported lower anxiety at the onset of treatment relative to patients with AN (reference group). Likewise, patients with OSFED reported significantly lower depression relative to patients with AN.

**Table 2.** Multi-level model results for each outcome. Sample n varies across analyses due to missing data.

| Variable | EDE-Q<br>(n<sub>obs</sub> = 4120; n<sub>patients</sub> = 1410) |  |  |  | Anxiety<br>(n<sub>obs</sub> = 4476; n<sub>patients</sub> = 1462) |  |  |  | Depression<br>(n<sub>obs</sub> = 4458; n<sub>patients</sub> = 1465) |  |  |  |
| --- | --- | --- | --- | --- | --- | --- | --- | --- | --- | --- | --- | --- |
|  | *b* (se) | Lower CI | Upper CI | *p* | *b* (se) | Lower CI | Upper CI | *p* | *b* (se) | Lower CI | Upper CI | *p* |
| **Fixed effects:** |  |  |  |  |  |  |  |  |  |  |  |  |
| Age | <.01 (<.01) | <.01 | 0.01 | .22 | -0.03 (0.01) | -0.06 | <.01 | .02 | -0.01 (0.01) | -0.04 | 0.02 | .45 |
| Treatment week | -0.41 (0.04) | -0.49 | -0.33 | <.001 | -0.86 (0.15) | -1.16 | -0.57 | <.001 | -0.86 (0.16) | -1.18 | -0.55 | <.001 |
| *Gender (Cis-gender female = 0)* |  |  |  |  |  |  |  |  |  |  |  |  |
| Cis-gender male | -0.26 (0.14) | -0.53 | 0.02 | .06 | -1.56 (0.58) | -2.69 | -0.42 | .01 | -1 (0.59) | -2.16 | 0.16 | .09 |
| Transgender/Non-binary/Other | 0.54 (0.16) | 0.22 | 0.85 | <.001 | 1.82 (0.66) | 0.52 | 3.12 | .01 | 3.04 (0.69) | 1.69 | 4.39 | <.001 |
| *Diagnosis (AN = Reference)* |  |  |  |  |  |  |  |  |  |  |  |  |
| BED | 0.08 (0.10) | -0.11 | 0.26 | .42 | -1.41 (0.4) | -2.19 | -0.63 | <.001 | 0.32 (0.41) | -0.49 | 1.12 | .44 |
| BN | 0.27 (0.14) | -0.02 | 0.55 | .07 | 0.44 (0.61) | -0.76 | 1.63 | .47 | 1 (0.63) | -0.22 | 2.23 | .11 |
| OSFED | 0.11 (0.12) | -0.12 | 0.35 | .33 | -0.08 (0.48) | -1.02 | 0.87 | .87 | 1.2 (0.5) | 0.23 | 2.17 | .02 |
| Age x Treatment week | <.01 (<.01) | <.01 | <.01 | .05 | 0.01 (0) | <.01 | 0.01 | .24 | 0 (0.01) | -0.01 | 0.01 | .46 |
| Cis-gender male x Treatment week | -0.09 (0.06) | -0.20 | 0.01 | .09 | -0.04 (0.2) | -0.42 | 0.35 | .86 | 0.07 (0.22) | -0.36 | 0.50 | .75 |
| Transgender/Non-binary/Other x Treatment week | -0.01 (0.06) | -0.13 | 0.11 | .86 | -0.1 (0.22) | -0.53 | 0.34 | .67 | -0.23 (0.25) | -0.71 | 0.25 | .35 |
| BED x Treatment week | -0.11 (0.04) | -0.18 | -0.04 | <.001 | -0.1 (0.13) | -0.37 | 0.16 | .44 | -0.59 (0.15) | -0.88 | -0.3 | <.001 |
| BN x Treatment week | -0.10 (0.06) | -0.22 | 0.01 | .06 | -0.11 (0.21) | -0.52 | 0.30 | .59 | -0.33 (0.23) | -0.78 | 0.13 | .16 |
| OSFED x Treatment week | -0.02 (0.04) | -0.11 | 0.07 | .66 | 0.13 (0.15) | -0.17 | 0.43 | .40 | -0.03 (0.17) | -0.36 | 0.30 | .86 |
| **Random effects:** |  |  |  |  |  |  |  |  |  |  |  |  |
| SD (Patient) | 1.18 |  |  |  | 4.84 |  |  |  | 5.02 |  |  |  |
| SD (Treatment Week) | 0.32 |  |  |  | 0.82 |  |  |  | 1.11 |  |  |  |

### Treatment outcomes

Treatment time (in weeks) was significantly associated with improved symptoms across all outcomes. Overall, mean EDE-Q scores decreased from 3.63 to 2.60 at week 20 and 2.37 at week 40; mean PHQ-8 scores decreased from 11.53 to 9.22 at week 20 and 8.71 at week 40; and mean GAD-7 scores decreased from 11.69 to 9.52 at week 20 and 9.05 at week 40. Moreover, symptom improvement was not significantly moderated by age or gender. In general, symptoms improved similarly across diagnoses (see Figure 1; underlying estimates by diagnosis are available in Supplement Table S2).

**Figure 1.**
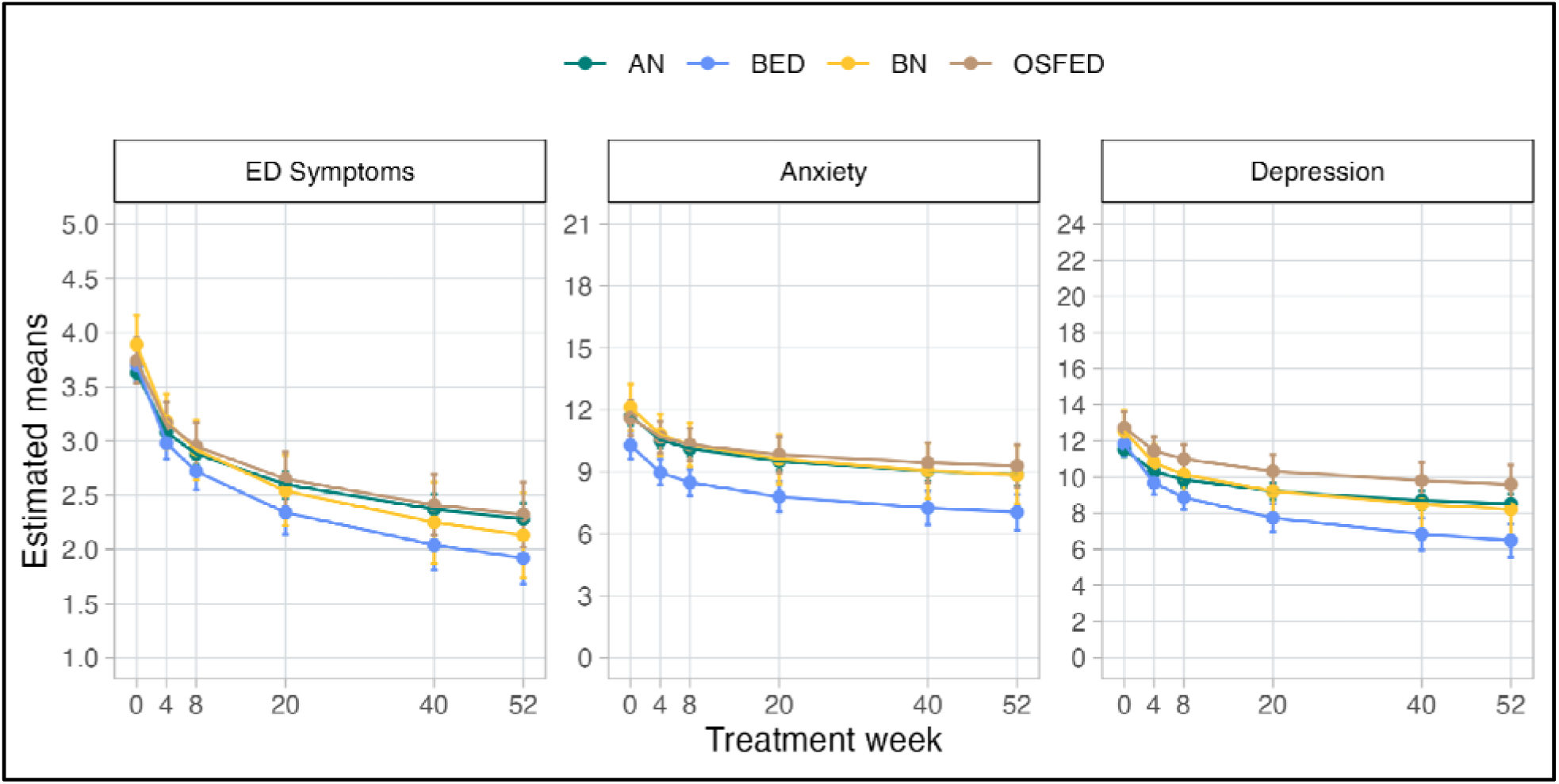
Model-derived estimates of outcomes over treatment time. AN = anorexia nervosa; BED = binge-eating disorder; BN = bulimia nervosa; OSFED = other specific eating or feeding disorder; ED = eating disorder.

Approximately 82% of patients entered treatment with clinically significant eating disorder symptoms (EDE-Q global score > 2.8), 63% of patients entered treatment with moderate-to-severe depression (PHQ-8 >= 10), and 56% with moderate anxiety (GAD-7 >= 10).

By 20 weeks of treatment, approximately 34% (CI: [31.06, 36.85]) of these patients showed improvement to sub-clinical levels of ED symptoms, 42% (CI: [38.63, 45.28]) for depression, and 40% (CI: [37.05, 43.87]) for anxiety. By week 40 of treatment, 49% (CI: [44.67, 53.19]) of patients decreased to subclinical levels on EDE-Q, 57.6% (CI: [52.7, 61.98] ) on depression, and 55.9% (CI: [50.84, 60.44]) on anxiety (see Figure 2).

**Figure 2.**
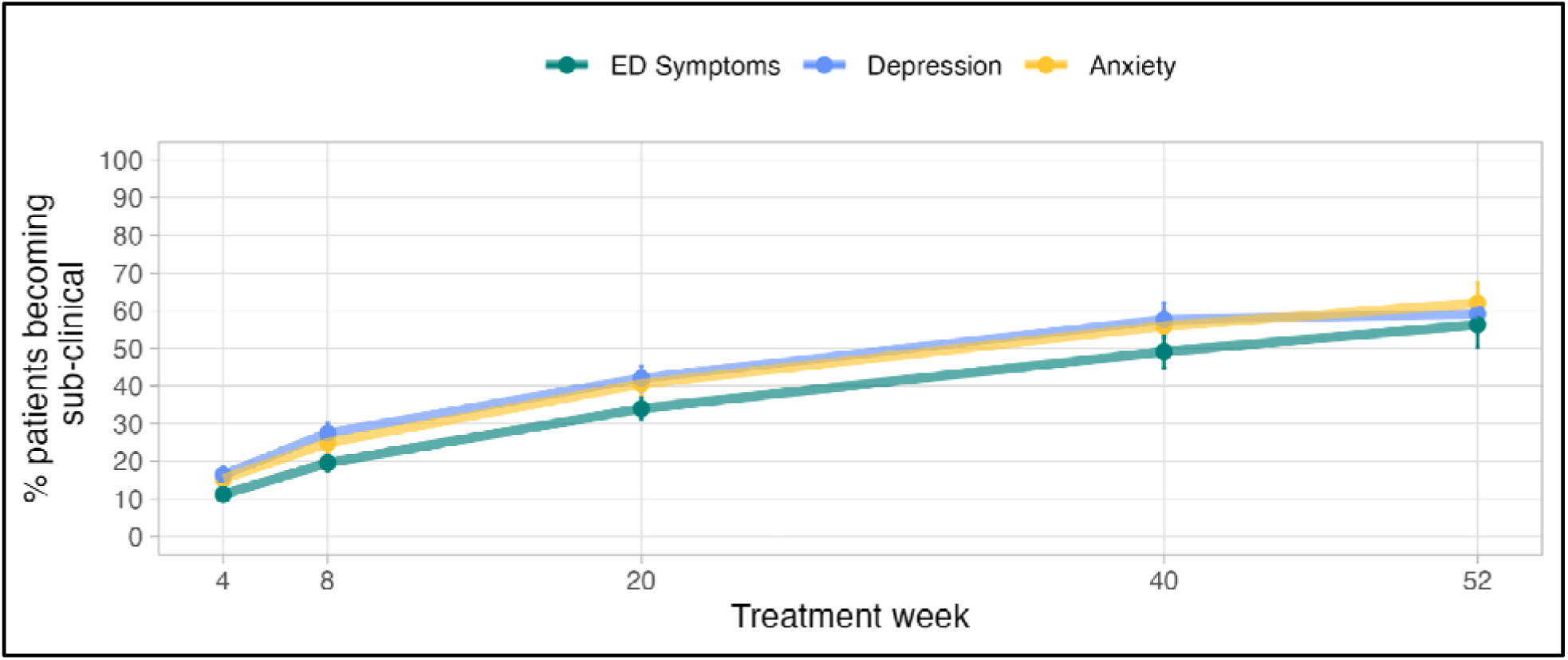
The percentage of patients who become subclinical on eating disorder symptoms, depression, and anxiety across treatment time. *Note:* Subclinical levels: anxiety (GAD-7 score < 10); depression (PHQ-8 score < 10); eating disorder (ED) symptoms (EDE-Q score < 2.8).

## Discussion

CBT-E delivered via telemedicine that includes a multidisciplinary treatment team is effective for improving eating disorder outcomes. Patients who received virtual CBT-E for up to one year showed significant improvements in weight restoration, eating disorder symptoms and comorbid depression and anxiety.

Results here show that CBT-E delivered virtually in an outpatient clinic setting by a multidisciplinary team produces clinically meaningful outcomes across eating disorder diagnoses. By week 40, more than half of patients requiring weight restoration were weight restored. Approximately half of patients who reported moderate-to-severe symptom scores on the EDE-Q, PHQ-8, and/or the GAD-7 at admission also showed a decrease in scores to a sub- clinical level by week 40. On average, symptom score averages also decreased from moderate-to-severe at admission to subclinical levels by week 20. These findings corroborate clinical trials (Fairburn et al., 2009; Fairburn et al, 2013), which generally show about 50% of patients have a BMI greater than 18.5 by end-of-treatment (for those that need to gain weight; however, this does not mean the patient reached their target weight) and 30% to 60% of patients with AN or BN are defined as in remission (definitions vary; Byrne et al., 2017; Groff, 2015; Fairburn et al., 2015; Byrne, Fursland, Allen, & Watson, 2011; de Jong et al., 2020). However, our findings show the complexities of translating results from controlled studies to a general patient population. For example, clinical trials of CBT-E typically have a fixed end point for treatment at approximately 20 or 40 weeks (depending on weight status). However, in our transdiagnostic sample of patients, the overall median length of treatment was 25 weeks [Q1: 9 weeks; Q3: 52 weeks]. Given the variability of real world treatment, it is challenging to make comparisons with clinical trial end-of-treatment outcomes.

Clinical trials have strict criteria for study inclusion, which may limit the generalizability of the results to the typical patient. For patients with eating disorders, comorbidity is the rule rather than the exception (Pearlstein, 2002), and clinical trials often exclude patients with certain comorbidities that are common in eating disorders (e.g., suicidality, substance use disorder, medical instability). The ability to access and join a clinical trial is also limited. Given these added challenges, the typical eating disorder patient may have additional barriers to success compared with those enrolled in a clinical trial. Even trials designed to be “inclusive” may not represent the average eating disorder patient (e.g., BMI criteria, no previous similar treatment, no psychiatric meds, non-US studies have universal healthcare, non-underweight, consent to research; Steinberg et al., 2023; Byrne, Fursland, Allen, & Watson, 2011). Therefore, it is important to consider outcomes within the context of such additional confounding factors that may have an impact on length of care and treatment outcomes.

The goal of this study was not to compare the effectiveness of CBT-E between eating disorder diagnoses; however, we report symptom outcomes by diagnosis for informational purposes. A similar trend was observed across all diagnoses for all symptoms such that self-report symptom scores continually and similarly decreased (i.e., symptom improvement) across time in treatment. Thus, our results provide evidence that CBT-E delivered virtually is a reasonable treatment approach for transdiagnostic eating disorders in an outpatient setting.

Although speculative, perhaps outpatient treatment delivered virtually for patients with an eating disorder specifically allows for greater patient retention and therefore improvement.

We acknowledge several limitations of this study. Our patient sample was somewhat homogenous due to self-selection in treatment. As we note earlier, the variability of time in treatment makes comparisons to clinical trials challenging; however, this is the reality of real-world treatment where patients leave treatment for a variety of reasons including financial, ambivalence toward recovery, or others. Moreover, the variability in treatment length for our patients makes it challenging to evaluate a clear end-of-treatment remission and/or recovery rate as an outcome. We also noted that missing data yielded different analytic samples across our analyses. However, we found that imputing the missing data led to nearly identical results.

Despite these limitations, this is the largest study to date evaluating the effectiveness of CBT-E for transdiagnostic eating disorders delivered in a virtual outpatient clinic setting by a multidisciplinary team. We used rigorous modeling techniques to derive estimates of treatment outcomes in this setting to show the effectiveness of CBT-E in this transdiagnostic sample. The additional implementation of virtual treatment for eating disorders will further advance access to care for those in need and only serve to improve overall patient outcomes.

## Data Availability

De-identified aggregate data is available upon reasonable request.

## Data Availability Statement

The data that support the findings of this study are derived from patient medical record data. The data are not publicly available and individual data cannot be shared due to privacy restrictions.

