## Supplemental table for "Treatment Outcome of Adults Receiving Virtual Cognitive Behavior Therapy for an Eating Disorder"

| **Table S1.** Percent of patients having each characteristic by diagnosis | | | | |
| --- | --- | --- | --- | --- |
| **Variable** | **AN** | **BED** | **BN** | **OSFED** |
| *Gender* |  |  |  |  |
| Cisgender female | 57.66 | 22.91 | 7.73 | 11.7 |
| Cisgender male | 47.41 | 39.26 | 7.41 | 5.93 |
| Transgender/Non-binary/Other | 67.78 | 15.56 | 1.11 | 15.56 |
| *Race/Ethnicity* |  |  |  |  |
| White | 59.45 | 22.73 | 6.74 | 11.07 |
| Multi-ethnic/racial | 56.8 | 21.3 | 10.65 | 11.24 |
| Asian | 63.1 | 15.48 | 7.14 | 14.29 |
| Hispanic | 51.02 | 29.59 | 8.16 | 11.22 |
| Black/African American | 42.65 | 36.76 | 5.88 | 14.71 |
| Other | 48.28 | 31.03 | 10.34 | 10.34 |
| *Prior Treatment* |  |  |  |  |
| None | 43.43 | 34.36 | 8.29 | 13.92 |
| Other | 59.42 | 26.09 | 7.25 | 7.25 |
| Prior HLOC | 81.38 | 4.66 | 6.28 | 7.69 |
| Unknown | 61.95 | 21.46 | 6.34 | 10.24 |

| **Table S2.** Weekly estimates for each outcome by diagnosis | | | | | | | |
| --- | --- | --- | --- | --- | --- | --- | --- |
| **Diagnosis** | **Outcome** | **Week 0** | **Week 4** | **Week 8** | **Week 20** | **Week 40** | **Week 52** |
| AN | ED Symptoms | 3.63 | 3.08 | 2.88 | 2.6 | 2.37 | 2.28 |
| BED | ED Symptoms | 3.7 | 2.98 | 2.72 | 2.34 | 2.04 | 1.92 |
| BN | ED Symptoms | 3.89 | 3.18 | 2.92 | 2.54 | 2.25 | 2.13 |
| OSFED | ED Symptoms | 3.74 | 3.16 | 2.95 | 2.65 | 2.41 | 2.32 |
| AN | Depression | 11.53 | 10.31 | 9.86 | 9.22 | 8.71 | 8.52 |
| BED | Depression | 11.85 | 9.68 | 8.88 | 7.74 | 6.84 | 6.49 |
| BN | Depression | 12.54 | 10.79 | 10.15 | 9.22 | 8.5 | 8.22 |
| OSFED | Depression | 12.73 | 11.46 | 11 | 10.33 | 9.81 | 9.6 |
| AN | Anxiety | 11.69 | 10.55 | 10.13 | 9.52 | 9.05 | 8.86 |
| BED | Anxiety | 10.29 | 8.97 | 8.49 | 7.8 | 7.26 | 7.05 |
| BN | Anxiety | 12.13 | 10.8 | 10.32 | 9.62 | 9.07 | 8.85 |
| OSFED | Anxiety | 11.62 | 10.68 | 10.33 | 9.84 | 9.45 | 9.3 |
| Patient Avg. | ED Symptoms | 3.63 | 3.08 | 2.88 | 2.6 | 2.37 | 2.28 |
| Patient Avg. | Depression | 11.53 | 10.31 | 9.86 | 9.22 | 8.71 | 8.52 |
| Patient Avg. | Anxiety | 11.69 | 10.55 | 10.13 | 9.52 | 9.05 | 8.86 |
